# Trends and determinants of minimum dietary diversity among children aged 6-23 months from 2014 to 2022 in Bangladesh: An analysis of nationally representative data

**DOI:** 10.64898/2026.04.13.26350794

**Authors:** Imteaz Mahmud, Mousumi Akter Mim, Kedir Teji Roba, Tanvir M Huda

## Abstract

**Introduction:** Minimum dietary diversity (MDD) is a key indicator of complementary feeding among children aged 6-23 months. This study examines the prevalence, trends, and determinants of MDD in Bangladesh over the period 2014 - 2022.

**Design:** Secondary analysis of the Bangladesh Demographic and Health Survey (BDHS) data between 2014 and 2022. The primary outcome was MDD defined as consumption of at least 5 of 8 food groups (MDD-8). We included 6,080 children aged 6–23 months to assess trends over time. The pooled datasets were used to identify factors associated with MDD-8. Multiple logistic regression was performed to assess the association between different factors and MDD-8, accounting for the complex survey design.

**Setting:** Bangladesh

**Results:** The proportion of children achieving MDD-8 increased from 26.4% in 2014 to 38.7% in 2017, but plateaued at 37.1% in 2022, with an average annual increase of 4.3% between 2014 and 2022. MDD-8 improved with child age. Higher odds of achieving MDD-8 were observed among children surveyed in later years, from wealthier households, with mothers who had ≥4 ANC visits, received PNC, had higher education, were employed, and had media exposure. Older age and higher birth order were also associated with achieving adequate MDD. Children in Chattogram and Sylhet were less likely to meet MDD-8 compared to Dhaka.

**Conclusions:** While dietary diversity improved between 2014 and 2017, progress stalled thereafter. Targeted, multisectoral strategies focusing on women’s empowerment, health service utilisation, media engagement, and disadvantaged regions are needed to improve child dietary diversity in Bangladesh.

## Introduction

Child malnutrition in all its forms remains a global health concern. According to the 2025 joint child malnutrition estimates, 150.2 million children under five are stunted, and 42.8 million are wasted (1). South Asia bears the highest burden of both wasting and stunting (1). In Bangladesh, 24% of children under 5 are stunted and 11% wasted (2) – levels classified as “high” by the WHO–UNICEF Technical Expert Advisory Group on Nutrition Monitoring (3). Achieving Sustainable Development Goal (SDG) 2.2, which targets to end all forms of malnutrition and a 40% reduction in the number of stunted children by 2030 (4), requires improvements in infant and young child feeding. Ensuring a diversified diet among children aged 6-23 months is critical to meeting these targets.

Adequate nutrition during the first 1,000 days is critical for optimal growth, cognitive development, and long-term health. Dietary diversity among children aged 6–23 months is a key indicator of complementary feeding quality and a proxy for dietary adequacy and household food security. It is widely used to monitor national nutrition progress and has been incorporated into SDG indicators (5). In 2008, the World Health Organisation (WHO) defined minimum dietary diversity (MDD-7) as consumption of at least 4 of 7 food groups (6). In 2017, this was revised to MDD-8 by adding breastmilk, requiring consumption of at least 5 of 8 food groups (7). While this improves measurement, it may result in lower prevalence estimates. Across LMICs, MDD-8 prevalence ranges from 18% in Sub-Saharan Africa to 54% in Latin America, with South Asia at 23.4% (8, 9). In Bangladesh, MDD-8 was reported at 38% in 2017–18 (8). However, existing studies have largely focused on MDD-7, and evidence on MDD-8 trends in Bangladesh remains limited.

Evidence suggests that maternal education, maternal employment, higher household wealth, exposure to mass media (including newspapers and magazines), attending more than four antenatal care (ANC) visits, and older child age are associated with greater dietary diversity in Bangladesh (8). However, there has been no recent analysis of the factors associated with MDD-8 in Bangladesh. Understanding dietary diversity and its trends is essential for assessing the current situation and the country’s progress in nutritional status. It is also important to generate up-to-date evidence on the newest indicators of dietary practices among Bangladeshi families. The performance of this new indicator may help evaluate the country’s progress towards achieving the SDG targets.

Therefore, this study aimed to examine the trends and determinants of MDD-8 among children aged 6–23 months using nationally representative BDHS data collected in 2014, 2017, and 2022, and to assess the association between MDD-8 and different forms of malnutrition.

## Methods

### 2.1 Study design

We analysed data from the Bangladesh Demographic and Health Surveys (BDHS) conducted between 2014 and 2022 (10). Since 1984, the DHS program has implemented standardised household surveys across multiple countries, generating reliable and comparable data on health and nutrition. In Bangladesh, these cross-sectional, nationally representative surveys have been conducted every 3 to 4 years since 1993, including the most recent survey in 2022. This study focused on children aged 6–23 months. We estimated the prevalence of MDD-8 in 2014, 2017, and 2022, examined factors associated with MDD-8, and assessed trends and consumption patterns across different food groups over time. This historical comparison enables an assessment of progress, or lack thereof, in addressing malnutrition among children aged 6–23 months in Bangladesh.

### 2.2 Sampling strategy and data collection

The BDHS used a two-stage cluster sampling design to obtain a nationally representative sample (2). The sampling frame was based on the most recent national census Enumeration Areas (EAs), each comprising approximately 120 households and serving as Primary Sampling Units (PSUs). In the first stage, EAs were stratified by urban and rural residence, and PSUs were selected using probability proportional to size. In the second stage, an average of 30 households per EA were randomly selected for data collection. Trained interviewers visited the selected households and collected demographic, household, maternal, paternal, and child-related information using the standard DHS questionnaires.

### 2.3 Variables

Children aged 6–23 months were identified from the children’s dataset. The primary outcome of interest was MDD-8, which was derived from children’s dietary intake information. In 2017, the WHO/UNICEF Technical Expert Advisory Group on Nutrition Monitoring (TEAM) recommended that consumption of at least 5 out of 8 food groups should be used to define MDD-8(7). In this analysis, we applied the MDD-8 criteria. Table 1 presents the breakdown of the food group components included in MDD-8 and MDD-7.

**Table 1.** Dietary diversity food group components.

| No | Food groups | Dietary diversity components |  |
| --- | --- | --- | --- |
|  |  | MDD-7* | MDD-8† |
| 1. | Grains, roots and tubers | ✓ | ✓ |
| 2. | Legumes and nuts | ✓ | ✓ |
| 3. | Dairy products | ✓ | ✓ |
| 4. | Flesh foods | ✓ | ✓ |
| 5. | Eggs | ✓ | ✓ |
| 6. | Vitamin A-rich fruits and vegetables | ✓ | ✓ |
| 7. | Other fruits and vegetables | ✓ | ✓ |
| 8. | Breastmilk |  | ✓ |

*MDD, minimum dietary diversity.*
*\*Based on the WHO indicator of MDD of at least four of the seven food groups in 2008(6).*
*†Based on the WHO indicator of MDD of at least five of eight food groups in 2017(7)*

To examine the determinants of MDD-8, we selected key child, maternal, paternal, and household factors based on the literature (11, 12). Variables were recoded according to established definitions. Child characteristics included age group (6–11, 12–17, and 18–23 months), sex (male or female), and birth order (firstborn, second-born, third-born, or higher). Maternal characteristics included current age (<20, 20–34, and ≥35 years), education level (none, primary, secondary, higher), employment status (working, not working), number of ANC visits (<4, ≥4), receipt of postnatal care (PNC) within 2 months of delivery (yes, no), and weekly media exposure (none, any media). Household characteristics included region (Dhaka, Barisal, Chattogram, Khulna, Mymensingh, Rajshahi, Rangpur, Sylhet), place of residence (urban or rural), sex of the household head (male, female), religion (Muslim, Hindu and/or others), number of children under 5 years in the household (≤2, >2), maternal involvement in household decision-making (involved, not involved), and wealth quintile (poorest, poorer, middle, richer, richest). Paternal characteristics included father’s education level (none, primary, secondary, higher). All variables were defined using standard DHS methodologies, with detailed descriptions available in the DHS manual (13). Household decision-making included decisions related to healthcare and major household purchases, considering whether mothers participated independently or jointly with their spouses. Weekly media exposure included reading newspapers or magazines, watching television, or listening to the radio.

### 2.4 Statistical analysis

Statistical analyses were performed using STATA 16 (14). Sampling weights were applied to estimate frequencies and percentages, accounting for under- and oversampling. To address the complex survey design, we used the svyset command in STATA to obtain robust standard errors. We estimated the prevalence of MDD-8 for the 2014, 2017, and 2022 surveys and calculated the average annual rate of change in overall and across subgroups defined by various variables. We also examined trends in the consumption of different food groups over the past decade and assessed changes in mean dietary diversity across the three survey rounds. The distribution of study variables was described for each survey year. Univariate logistic regression was used to assess the association between each determinant and MDD-8, reported as unadjusted odds ratios (ORs). Multivariable logistic regression was then conducted to examine adjusted associations between covariates and MDD-8. Survey year was included in the adjusted model, and potential interactions between variables were also explored.

### 2.5 Ethical considerations

This study used publicly available, de-identified BDHS data obtained with permission from the DHS Program. The BDHS surveys were approved by the Institutional Review Board (IRB) of ICF International and the Bangladesh Medical Research Council (BMRC). As the dataset was anonymised, no additional ethical approval was required.

## 3. Results

### 3.1 Participants

Between 2014 and 2022, a total of 6,080 children aged 6–23 months were included. The distribution of the sample across the eight divisions of Bangladesh is presented in Table 2. Most participants were from Dhaka and Chattogram divisions, while Sylhet had the lowest representation (8.4%).

**Table 2:** Distribution of the sample population in eight districts of Bangladesh.

| Division | Year 2014 - 2022 |  |
| --- | --- | --- |
|  | Frequency | Percentage |
| Barisal | 662 | 10.9 |
| Chattogram | 1,059 | 17.4 |
| Dhaka | 991 | 16.3 |
| Khulna | 657 | 10.8 |
| Mymensingh | 759 | 12.5 |
| Rajshahi | 665 | 10.9 |
| Rangpur | 778 | 12.8 |
| Sylhet | 509 | 8.4 |
| Total | 6,080 | 100 |

### 3.2 Trends in minimum dietary diversity and child characteristics

Table 3 shows the distribution of MDD-8 across subpopulations over time. Overall, the proportion of children receiving a diversified diet increased from 26.4% (95% CI: 24.3%, 28.7%) in 2014 to 38.7% (95% CI: 36.5%, 40.0%) in 2017, followed by a slight decline to 37.1% (95% CI: 33.9%, 40.5%) in 2022. Improvements were observed across most subgroups between 2014 and 2017, with a modest decline in 2022. The average annual rate of change (AARC) from 2014 to 2022 was 4.3%.

**Table 3:** The proportion of minimum dietary diversity (MDD-8*) among children aged 6–23 months in Bangladesh from 2014 to 2022.

| Characteristics | Category | Year |  |  |  |  |  |  |  |  | AARC<sup>†</sup> |
| --- | --- | --- | --- | --- | --- | --- | --- | --- | --- | --- | --- |
|  |  | 2014 |  |  | 2017 |  |  | 2022 |  |  | 2014 - 2022 |
|  |  | n | % | 95% CI | n | % | 95% CI | n | % | 95% CI | % |
| **Overall** |  | 648 | 26.4 | 24.3 – 28.7 | 970 | 38.7 | 36.5 – 40.0 | 475 | 37.1 | 33.9 – 40.5 | 4.3 |
| **Children Characteristics** |  |  |  |  |  |  |  |  |  |  |  |
| Children age category | 6–11 months | 131 | 13.7 | 11.3 - 16.6 | 204 | 23.8 | 20.6 - 27.3 | 118 | 24.1 | 19.9 - 28.9 | 7.3 |
|  | 12–17 months | 249 | 29.6 | 26.0 - 33.4 | 386 | 45.9 | 42.1 - 49.7 | 154 | 42.3 | 36.4 - 48.4 | 4.5 |
|  | 18–23 months | 268 | 38.1 | 33.9 - 42.5 | 380 | 46.4 | 42.5 - 50.2 | 203 | 46.4 | 40.9 - 52.1 | 2.5 |
| Children sex | Male | 318 | 25.7 | 22.9 - 28.8 | 496 | 38.7 | 35.8 - 41.7 | 243 | 38.0 | 33.8 - 42.4 | 5.0 |
|  | Female | 330 | 27.2 | 24.1 - 30.6 | 474 | 38.8 | 35.6 - 42.1 | 232 | 36.3 | 31.8 - 40.9 | 3.7 |
| Birth order | Firstborn | 309 | 31.2 | 27.9 - 34.8 | 421 | 43.9 | 40.3 - 47.5 | 211 | 42.4 | 37.3 - 47.6 | 3.9 |
|  | Secondborn | 183 | 24 | 20.4 - 28.2 | 310 | 37.4 | 33.8 - 41.2 | 158 | 34.9 | 29.9 - 40.2 | 4.8 |
|  | Thirdborn or higher | 156 | 22.2 | 18.3 - 26.6 | 239 | 33.7 | 29.7 - 38.0 | 106 | 32.2 | 26.7 - 38.3 | 4.8 |
| **Maternal Characteristics** |  |  |  |  |  |  |  |  |  |  |  |
| Mother's age (current) | < 20 years | 199 | 26.7 | 22.9 - 31.0 | 279 | 39.9 | 35.8 - 44.2 | 110 | 36.4 | 30.6 - 42.7 | 3.9 |
|  | 20–34 years | 425 | 26.1 | 23.2 - 29.2 | 653 | 38.2 | 35.4 - 41.1 | 344 | 37.8 | 34.0 - 41.8 | 4.7 |
|  | ≥35 years | 24 | 30.8 | 19.9 - 44.3 | 38 | 39.8 | 29.6 - 50.8 | 21 | 30.8 | 19.4 - 45.2 | 0.00 |
| Mother's level of education | None | 42 | 13.5 | 9.3 - 19.3 | 31 | 18.6 | 12.7 - 26.3 | 16 | 26.5 | 16.6 - 39.5 | 8.8 |
|  | Primary | 124 | 18.1 | 14.8 - 21.9 | 194 | 30.8 | 26.8 - 35.1 | 65 | 23.1 | 18.0 - 29.0 | 3.1 |
|  | Secondary | 354 | 30.5 | 27.5 - 33.7 | 472 | 39.2 | 36.2 - 42.2 | 260 | 38.4 | 33.9 - 43.0 | 2.9 |
|  | Higher | 118 | 44.8 | 37.9 - 51.9 | 273 | 56.7 | 51.4 - 61.9 | 134 | 53.4 | 46.4 - 60.3 | 2.2 |
| Mother's working status | Working | 142 | 26.9 | 22.2 - 32.1 | 360 | 38.3 | 35.3 - 41.3 | 114 | 40.6 | 34.2 - 47.4 | 5.3 |
|  | Not working | 505 | 26.3 | 24.0 - 28.8 | 610 | 39.7 | 36.2 - 43.3 | 361 | 36.1 | 32.5 - 39.9 | 4.0 |
| Mother's access to media at least once a week | None | 177 | 17.4 | 14.5 - 20.9 | 257 | 29.5 | 25.9 - 33.4 | 158 | 28.0 | 24.0 - 32.4 | 6.1 |
|  | Any media | 471 | 31.9 | 29.0 - 34.9 | 713 | 43.5 | 40.8 - 46.3 | 317 | 44.5 | 39.7 - 49.4 | 4.2 |
| Number of antenatal care visits | < 4 | 381 | 23.5 | 20.8 - 26.3 | 382 | 30.1 | 27.3 - 33.1 | 235 | 32.1 | 28.2 - 36.2 | 3.9 |
|  | ≥ 4 | 267 | 33.8 | 29.4 - 38.6 | 583 | 50 | 46.6 - 53.4 | 239 | 46.3 | 41.2 - 51.5 | 4.0 |
| Postnatal check within 2 months | Yes | 457 | 30.6 | 27.8 - 33.5 | 665 | 41.3 | 38.7 - 44.0 | 193 | 38.0 | 33.3 - 43.0 | 2.7 |
|  | No | 191 | 20 | 16.3 - 24.3 | 298 | 35.5 | 31.9 - 39.2 | 281 | 37.7 | 33.5 - 42.0 | 8.2 |
| **Paternal characteristics** |  |  |  |  |  |  |  |  |  |  |  |
| Father's level of education | None | 98 | 17.9 | 14.1 - 22.5 | 88 | 27.5 | 22.3 - 33.3 | 45 | 26.5 | 19.3 - 35.1 | 5.0 |
|  | Primary | 173 | 24.3 | 20.0 - 29.1 | 283 | 32.8 | 29.3 - 36.5 | 116 | 29.6 | 24.7 - 35.1 | 2.5 |
|  | Secondary | 228 | 27.5 | 24.0 - 31.3 | 309 | 39.7 | 36.0 - 43.6 | 173 | 41.3 | 36.0 - 46.8 | 5.2 |
|  | Higher | 148 | 41 | 35.8 - 46.4 | 281 | 56.5 | 51.5 - 61.3 | 139 | 50.8 | 43.7 - 58.0 | 2.7 |
| **Household Characteristics** |  |  |  |  |  |  |  |  |  |  |  |
| Residence | Urban | 249 | 33.2 | 29.1 - 37.5 | 382 | 46 | 41.5 - 50.5 | 169 | 44.5 | 38.2 - 51.1 | 3.7 |
|  | Rural | 399 | 24 | 21.5 - 26.7 | 588 | 36.2 | 33.7 - 38.8 | 306 | 34.5 | 30.7 - 38.4 | 4.6 |
| Region | Dhaka | 124 | 26.9 | 23.2 - 31.0 | 149 | 41 | 35.7 - 46.4 | 91 | 46.1 | 38.4 - 54.1 | 6.9 |
|  | Barishal | 76 | 28.4 | 22.7 - 34.9 | 94 | 34.2 | 28.2 - 40.9 | 36 | 26.5 | 19.3 - 35.2 | -0.8 |
|  | Chattogram | 110 | 22.1 | 17.3 - 27.9 | 160 | 39.3 | 34.1 - 44.7 | 56 | 23.3 | 16.8 - 31.3 | 0.6 |
|  | Khulna | 92 | 34.8 | 28.9 - 41.2 | 97 | 38 | 32.2 - 44.2 | 75 | 54.1 | 47.2 - 60.8 | 5.7 |
|  | Mymensingh | - | - | - | 109 | 36.5 | 29.1 - 44.7 | 69 | 41.9 | 33.6 - 50.7 | 2.8 |
|  | Rajshahi | 83 | 26.6 | 20.8 - 33.4 | 100 | 35.1 | 29.3 - 41.5 | 56 | 43.3 | 33.2 - 53.9 | 6.3 |
|  | Rangpur | 91 | 28.1 | 22.4 - 34.6 | 139 | 46.8 | 41.3 - 52.4 | 58 | 35.1 | 27.2 - 44.0 | 2.8 |
|  | Sylhet | 72 | 24 | 17.1 - 32.5 | 122 | 31.1 | 25.8 - 36.9 | 34 | 21.9 | 15.9 - 29.4 | -1.1 |
| Wealth Index | Poorest | 87 | 15.7 | 11.8 - 20.6 | 137 | 26.3 | 22.2 - 30.9 | 69 | 27.1 | 21.7 - 33.2 | 7.1 |
|  | Poorer | 107 | 24 | 19.7 - 28.9 | 179 | 33.9 | 29.5 - 38.7 | 91 | 34.1 | 27.5 - 41.3 | 4.5 |
|  | Middle | 119 | 25.7 | 21.5 - 30.4 | 160 | 37.5 | 32.6 - 42.6 | 81 | 32.1 | 25.9 - 39.0 | 2.8 |
|  | Richer | 168 | 30.7 | 26.0 - 35.9 | 213 | 41.2 | 36.7 - 45.9 | 104 | 41.4 | 34.8 - 48.3 | 3.8 |
|  | Richest | 167 | 36.6 | 31.2 - 42.3 | 281 | 56.3 | 51.0 - 61.4 | 130 | 53.6 | 45.9 - 61.1 | 4.9 |
| Sex of the Household head | Male | 588 | 26.5 | 24.2 - 28.8 | 838 | 38.6 | 36.2 - 41.0 | 420 | 37.2 | 33.7 - 40.9 | 4.3 |
|  | Female | 60 | 25.9 | 19.0 - 34.3 | 132 | 39.8 | 33.9 - 45.9 | 55 | 36.7 | 28.2 - 46.2 | 4.4 |
| Religion | Muslims | 592 | 26.6 | 24.3 - 28.9 | 902 | 39.2 | 36.9 - 41.5 | 430 | 37.0 | 33.5 - 40.5 | 4.2 |
|  | Hindus and others | 56 | 24.9 | 16.0 - 36.6 | 68 | 33 | 25.1 - 42.0 | 45 | 39.4 | 28.5 - 51.4 | 5.9 |
| Number of children under 5 years of age within the household | <=2 | 623 | 26.8 | 24.6 - 29.1 | 932 | 39.2 | 36.9 - 41.5 | 457 | 37.8 | 34.4 - 41.2 | 4.4 |
|  | >2 | 25 | 19.7 | 12.0 - 30.7 | 38 | 30.6 | 22.1 - 40.8 | 18 | 25.1 | 14.9 - 39.2 | 3.1 |
| Mother's involvement in household decision making | No | 194 | 26.4 | 22.7 - 30.6 | 183 | 39.5 | 34.7 - 44.6 | 41 | 43.6 | 32.9 - 54.9 | 6.5 |
|  | Yes | 296 | 25.4 | 22.4 - 28.5 | 576 | 39.7 | 36.6 - 42.8 | 292 | 35.5 | 31.7 - 39.6 | 4.3 |

*n*: weighted counts, % (95 % CI): weighted proportion and CI.
\*Based on the WHO indicator of MDD of at least five of the eight food groups in 2017(7)
† AARC = Average annual rate of change

MDD-8 increased steadily among children in Dhaka, Khulna, and Rajshahi divisions; those from the poorest households; younger children (6–11 months); non-Muslims; and children whose mothers had no education, were not involved in decision-making, had fewer than four ANC visits, no PNC, or were employed. Increased MDD-8 was also observed among children whose fathers had secondary education.

Figure 1 illustrates the trend of each food group’s consumption between 2014 and 2022.

**Figure 1:**
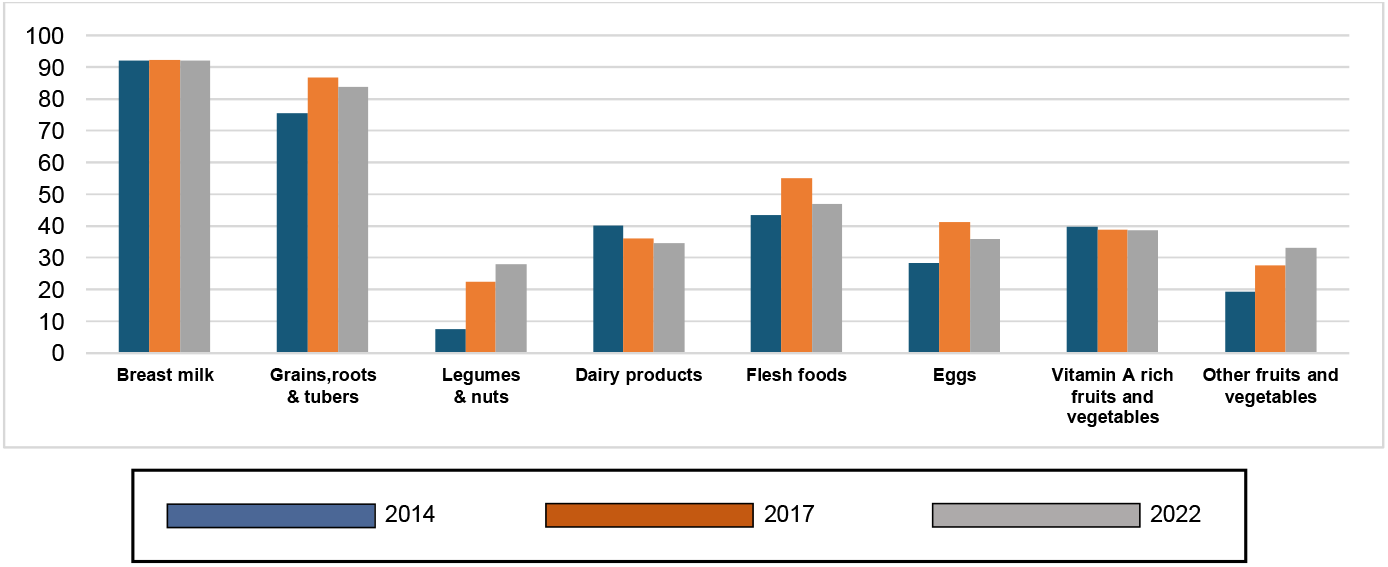
Consumption of various food groups between 2014 and 2022 among children aged 6-23 months.

Consumption of fruits, vegetables, legumes, and nuts increased over time, although intake of vitamin A-rich fruits and vegetables remained unchanged between 2017 and 2022. Breast milk consumption remained consistently high, while dairy consumption declined from 2014 to 2022. Intake of eggs, flesh foods, and staple foods increased between 2014 and 2017 but declined in 2022.

Figure 2 shows dietary diversity scores by child age. Overall, dietary diversity increased with age, except for a decline at 23 months in 2014.

**Figure 2:**
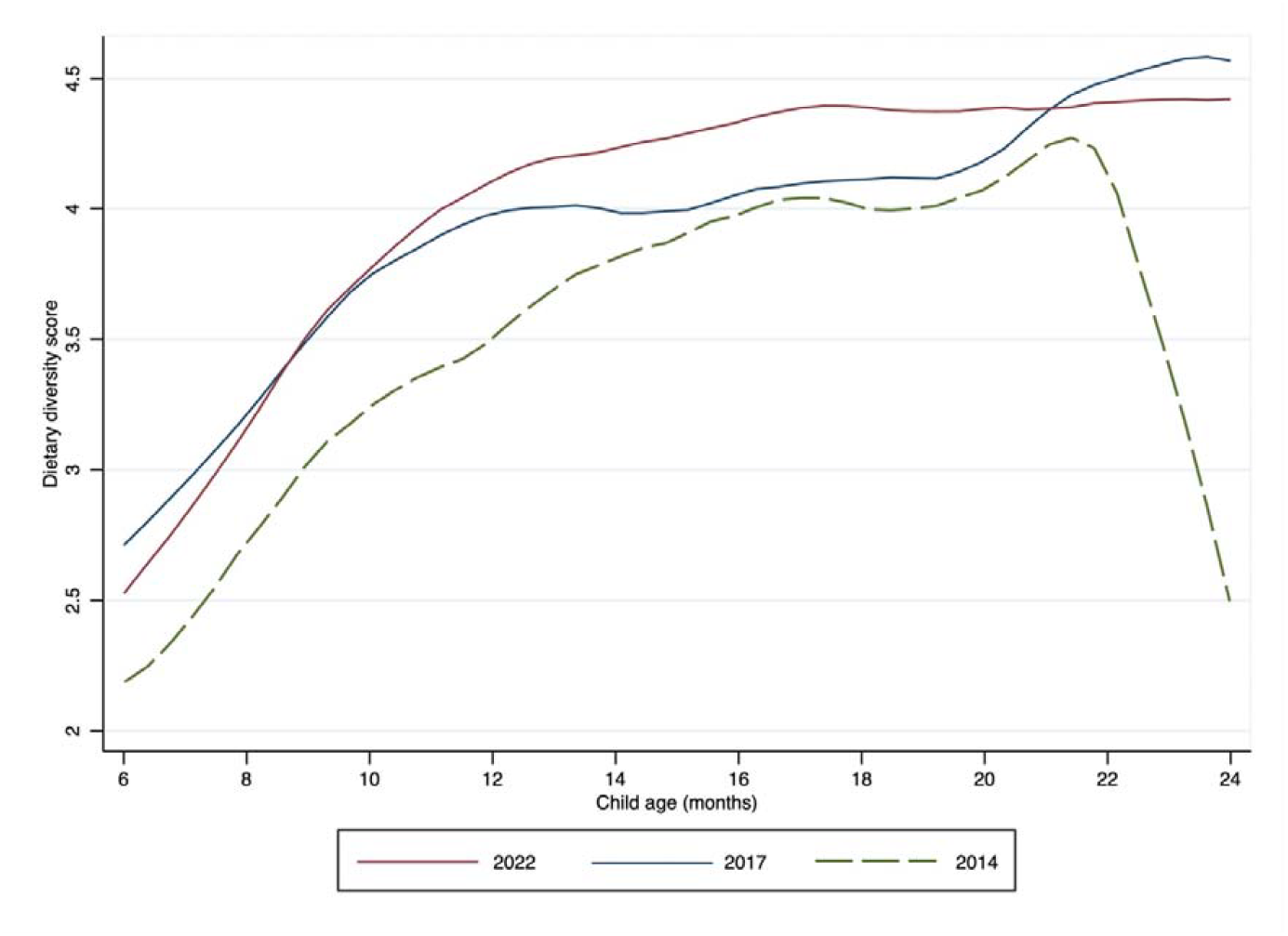
Mean dietary diversity scores by the child’s age in months.

The 2014 survey recorded the lowest average scores. In both 2017 and 2022, scores rose to around 4 by 12 months. The 2022 trend increased steadily until 17 months before plateauing, whereas the 2017 trend plateaued earlier but rose sharply after 19 months.

### 3.3 Factors associated with minimum dietary diversity

Table 4 presents unadjusted and adjusted odds ratios for MDD-8 determinants using pooled data from 2014–2022. Compared with 2014, children in 2017 had higher odds of achieving MDD-8. A clear dose–response relationship was observed with child age and maternal education, with the highest odds among children aged 18–23 months and those whose mothers had higher education. Children from the richest households had higher odds of MDD-8 compared with those in the middle quintile. Higher odds were also observed among children whose mothers attended ≥4 ANC visits, received PNC, were employed, or had media exposure. Children from Chattogram and Sylhet divisions had higher odds than those from Dhaka. In contrast, second- and third-born children had lower odds than first-borns.

**Table 4:** Factors associated with minimum dietary diversity (MDD-8*) among children aged 6–23 months in Bangladesh, showing unadjusted and adjusted OR from 2014 to 2022.

| Characteristics | Category | Year (2014 – 2022) |  |  |  |  |  |
| --- | --- | --- | --- | --- | --- | --- | --- |
|  |  | OR | 95% CI | P-value | AOR | 95% CI | P-value |
| Survey year | 2014 | Ref. |  |  | Ref. |  |  |
|  | 2017 | 1.76 | 1.52 - 2.05 | <0.001 | 1.8 | 1.49 - 2.17 | <0.001 |
|  | 2022 | 1.64 | 1.37 - 1.98 | <0.001 | 1.71 | 1.36 - 2.15 | <0.001 |
| **Children Characteristics** |  |  |  |  |  |  |  |
| Children age category | 6–11 months | Ref. |  |  | Ref. |  |  |
|  | 12–17 months | 2.57 | 2.19 - 3.01 | <0.001 | 2.68 | 2.21 - 3.25 | <0.001 |
|  | 18–23 months | 3.09 | 2.64 - 3.62 | <0.001 | 3.44 | 2.84 - 4.17 | <0.001 |
| Children sex | Male | Ref. |  |  | Ref. |  |  |
|  | Female | 1.01 | 0.9 - 1.14 | 0.80 | 0.93 | 0.80 - 1.08 | 0.33 |
| Birth order | Firstborn | Ref. |  |  | Ref. |  |  |
|  | Secondborn | 0.75 | 0.65 - 0.87 | <0.001 | 0.78 | 0.64 - 0.96 | 0.02 |
|  | Thirdborn or higher | 0.65 | 0.55 - 0.76 | <0.001 | 0.82 | 0.63 - 1.06 | 0.14 |
| **Maternal Characteristics** |  |  |  |  |  |  |  |
| Mother's age (current) | < 25 years | 1 | 0.87 - 1.15 | 0.98 | 0.93 | 0.76 - 1.14 | 0.48 |
|  | 25–34 years | Ref. |  |  | Ref. |  |  |
|  | >=35 years | 1.07 | 0.82 - 1.41 | 0.59 | 1.18 | 0.85 - 1.64 | 0.30 |
| Mother's level of education | None | Ref. |  |  | Ref. |  |  |
|  | Primary | 1.6 | 1.18 - 2.18 | 0.002 | 1.38 | 0.97 - 1.97 | 0.07 |
|  | Secondary | 2.77 | 2.08 - 3.71 | <0.001 | 2.01 | 1.38 - 2.93 | <0.001 |
|  | Higher | 5.6 | 4.09 - 7.65 | <0.001 | 2.86 | 1.84 - 4.46 | <0.001 |
| Mother's working status | Not working | Ref. |  |  | Ref. |  |  |
|  | Working outside home | 1.15 | 1.00 - 1.32 | 0.04 | 1.2 | 1.00 - 1.44 | 0.05 |
| Mother's access to media at least once a week | None | Ref. |  |  |  |  |  |
|  | Any media | 1.99 | 1.72 - 2.31 | <0.001 | 1.35 | 1.11 - 1.64 | 0.003 |
| Number of antenatal care visits | < 4 | Ref. |  |  | Ref. |  |  |
|  | ≥ 4 | 2.07 | 1.79 - 2.38 | <0.001 | 1.34 | 1.12 - 1.60 | 0.001 |
| Postnatal check within 2 months | No | Ref. |  |  | Ref. |  |  |
|  | Yes | 1.28 | 1.11 - 1.47 | 0.001 | 1.27 | 1.07 - 1.50 | 0.006 |
| **Paternal characteristics** |  |  |  |  |  |  |  |
| Father's level of education | None | Ref. |  |  | Ref. |  |  |
|  | Primary | 1.41 | 1.13 - 1.77 | 0.003 | 1.08 | 0.83 - 1.39 | 0.56 |
|  | Secondary | 1.87 | 1.53 - 2.30 | <0.001 | 1.03 | 0.79 - 1.35 | 0.80 |
|  | Higher | 3.44 | 2.75 - 4.32 | <0.001 | 1.26 | 0.91 - 1.75 | 0.15 |
| **Household Characteristics** |  |  |  |  |  |  |  |
| Residence | Urban | Ref. |  |  | Ref. |  |  |
|  | Rural | 0.66 | 0.57 - 0.76 | <0.001 | 0.93 | 0.77 - 1.12 | 0.44 |
| Region | Dhaka | Ref. |  |  | Ref. |  |  |
|  | Barishal | 0.8 | 0.63 - 1.02 | 0.07 | 1.03 | 0.77 - 1.37 | 0.83 |
|  | Chattogram | 0.75 | 0.60 - 0.94 | 0.01 | 0.73 | 0.57 - 0.93 | 0.01 |
|  | Khulna | 1.27 | 1.02 - 1.57 | 0.03 | 1.15 | 0.87 - 1.53 | 0.30 |
|  | Mymensingh | 0.92 | 0.72 - 1.18 | 0.53 | 1.09 | 0.83 - 1.45 | 0.52 |
|  | Rajshahi | 0.96 | 0.76 - 1.22 | 0.77 | 0.88 | 0.68 - 1.14 | 0.34 |
|  | Rangpur | 1.07 | 0.85 - 1.34 | 0.56 | 1.21 | 0.89 - 1.65 | 0.22 |
|  | Sylhet | 0.73 | 0.56 - 0.94 | 0.56 | 0.68 | 0.49 - 0.94 | 0.02 |
| Wealth Index | Poorest | 0.62 | 0.50 - 0.77 | <0.001 | 0.78 | 0.59 - 1.03 | 0.08 |
|  | Poorer | 0.95 | 0.78 - 1.16 | 0.61 | 1.1 | 0.86 - 1.40 | 0.44 |
|  | Middle | Ref. |  |  | Ref. |  |  |
|  | Richer | 1.28 | 1.06 - 1.54 | 0.01 | 1.19 | 0.94 - 1.50 | 0.14 |
|  | Richest | 1.99 | 1.62 - 2.43 | <0.001 | 1.45 | 1.09 - 1.92 | 0.01 |
| Sex of the Household head | Male | Ref. |  |  | Ref. |  |  |
|  | Female | 1.07 | 0.88 - 1.30 | 0.50 | 1.02 | 0.81 - 1.29 | 0.86 |
| Religion | Muslims | Ref. |  |  | Ref. |  |  |
|  | Hindus and others | 0.87 | 0.65 - 1.18 | 0.37 | 0.94 | 0.69 - 1.29 | 0.71 |
| Number of children under 5 years of age within the household | <=2 | Ref. |  |  | Ref. |  |  |
|  | >2 | 0.64 | 0.46 - 0.89 | 0.007 | 0.82 | 0.55 - 1.23 | 0.34 |
| Mother's involvement in household decision making | No | Ref. |  |  | Ref. |  |  |
|  | Yes | 1.05 | 0.90 - 1.23 | 0.49 | 0.89 | 0.75 - 1.05 | 0.17 |

OR, unadjusted Odds Ratio; AOR, adjusted Odds Ratio; CI, Confidence Interval; Ref, reference; P, P-value.
All variables included in the univariate analysis were also included in the multivariable analysis.
\*Based on the WHO indicator of MDD of at least five of the eight food groups in 2017(7)

No significant associations were found with child sex, sex of household head, place of residence, religion, number of under-5 children, maternal age, maternal decision-making, or paternal education.

## 4. Discussion

In this study, the proportion of children aged 6–23 months meeting MDD-8 increased from 26.4% in 2014 to 38.7% in 2017, before slightly declining to 37.1% in 2022. Over the past decade, consumption of legumes, nuts, fruits, and vegetables increased, although intake of vitamin A-rich fruits and vegetables remained unchanged between the two most recent surveys. Breastfeeding remained consistently high, whereas dairy consumption declined steadily. Consumption of flesh foods and grains, roots, and tubers increased from 2014 to 2017 but declined in 2022. In the pooled analysis, survey year, region, wealth quintile, ANC visits, PNC, maternal education, maternal employment, media exposure, child age, and birth order were associated with MDD-8. Although dietary diversity improved over time, the overall level remained low.

This study included 6,080 children aged 6–23 months from the 2014, 2017, and 2022 BDHS surveys. The highest prevalence of MDD-8 was observed in 2017 (38.7%), representing a marked improvement from 2014 (26.4%). Although the prevalence remained relatively high in 2022 (37.1%), the slight decline from 2017 suggests that progress may have stalled. Bangladesh’s 2022 estimate is higher than the average reported for South Asia (23.37%)(8), although it remains lower than that of countries such as the Maldives and Nepal (8). The improvement between 2014 and 2017 may reflect large-scale infant and young child feeding (IYCF) behaviour change programmes, including counselling and media-based interventions, together with improved integration of nutrition counselling into primary healthcare and broader socioeconomic progress (15-17). The slight decline in 2022 may partly reflect the effects of the COVID-19 pandemic, which disrupted livelihoods, food systems, and household purchasing power, thereby reducing access to nutritious foods(18, 19).

The food consumption patterns observed highlight both strengths and persistent gaps in complementary feeding practices in Bangladesh. Breastfeeding remained consistently high, likely reflecting strong cultural norms, national policy support such as the Breast-milk Substitutes Act 2013, the national IYCF strategy, and WHO recommendations to continue breastfeeding up to 2 years of age (20-22). High consumption of grains, roots, and tubers reflects the dominance of rice-based staples in the Bangladeshi diet. Earlier studies have shown that khichuri and suji are commonly used complementary foods in early infancy (23). In contrast, consumption of nutrient-dense food groups, including legumes and nuts, dairy products, eggs, and other fruits and vegetables, remained suboptimal. This may reflect affordability constraints, cultural feeding practices, and caregivers’ perceptions of age-appropriate foods. Consumption of vitamin A-rich fruits and vegetables was relatively higher, possibly due to the seasonal availability of mangoes, papayas, jackfruit, and pumpkins and the widespread practice of homestead gardening (24, 25). Moderate consumption of flesh foods may reflect the availability of fish in Bangladesh, although cost likely remains a barrier to regular intake of animal-source foods (26-28).

Consistent with previous studies, child age was strongly associated with MDD-8 (29-31). Older children may be more likely to consume diverse foods because they are more accustomed to different tastes and textures and increasingly share family foods (32). In Bangladesh, complementary feeding typically begins at 6 months with rice-based foods such as porridge or khichuri, followed later by foods from the family pot. Although mean dietary diversity increased with age across all survey years, a sharp decline at 23 months was observed in the 2014 survey. As the sample size in this age group was comparable with that of other age groups, this finding is unlikely to be due to small numbers. It may instead reflect a measurement artefact, such as age classification issues or incomplete recording of food group items in this subgroup (33).

Children of second or higher birth order had lower odds of meeting MDD-8 than first-born children, which is consistent with previous research (34, 35). This may reflect dilution of household resources, including money, time, and caregiver attention, as family size increases. Earlier BDHS analyses have similarly shown a nutritional disadvantage among later-born children (36). Becker’s quantity–quality trade-off model provides a useful explanation, suggesting that parental investment per child may decline as the number of children increases (37).

Maternal education was positively associated with MDD-8, in line with findings from Bangladesh and other low- and middle-income countries (8, 29-31, 38). More educated mothers may have better nutrition knowledge, greater exposure to dietary counselling, and stronger decision-making capacity, all of which may support healthier feeding practices. Media exposure was also positively associated with dietary diversity. Mothers exposed to newspapers, magazines, radio, or television at least once a week had greater odds of meeting MDD-8, a pattern also reported elsewhere (35, 39). Media can serve as an important source of health and nutrition information, and in Bangladesh, it has been used extensively to promote food security and balanced diets through national nutrition strategies and campaigns (40). However, some studies have reported negative associations, possibly due to reduced caregiver–child interaction or increased exposure to marketing of energy-dense, nutrient-poor foods (41, 42).

Receipt of maternal health services was another important predictor. Mothers who received PNC had higher odds of providing a diversified diet to their children, consistent with previous studies in Bangladesh and other settings (43, 44). PNC contacts may provide an opportunity for counselling on breastfeeding, complementary feeding, and age-appropriate foods. Similarly, mothers who attended four or more ANC visits had higher odds of achieving MDD-8 (43), likely reflecting both greater health service contact and better exposure to nutrition information.Maternal employment was positively associated with MDD-8. Similar findings have been reported in other countries, including Ghana and several South Asian settings (8, 44). Employment may improve household income, purchasing power, women’s empowerment, and decision-making capacity, thereby facilitating access to diverse foods(45). However, the relationship is complex. Time constraints associated with maternal employment may reduce childcare and feeding time, particularly in precarious jobs, as shown in Ethiopia and Ghana (46, 47). This suggests that the nutritional effects of maternal employment may depend on the type and conditions of work.

Household wealth showed a strong association with dietary diversity. Children from the richest households had significantly higher odds of meeting MDD-8 than those from middle-income households. This is consistent with evidence from South Asia, Southeast Asia, and rural Bangladesh, where greater wealth has been associated with better dietary diversity (8, 11, 48). Wealthier families are better able to afford diverse, nutrient-dense foods and may also have greater access to information and services that support optimal feeding practices.

Regional disparities were also evident. Children from Chattogram and Sylhet had higher odds of meeting MDD-8 than those from Dhaka. Previous studies in Bangladesh have also reported geographical variation in complementary feeding practices (49-51). These differences may reflect variation in food environments, agricultural production, climatic conditions, household resources, and cultural norms.

### 4.1 Strengths and limitations

This study has several limitations. As an observational cross-sectional analysis, it is vulnerable to confounding and cannot establish causality. The associations identified should therefore be interpreted as descriptive and hypothesis-generating. Recall bias and social desirability bias may have affected mothers’ reporting of child feeding practices. Measurement error may also have occurred despite standardised DHS training procedures. Selection bias is another consideration, as the children’s dataset included only children whose mothers were present and eligible for interview. Children whose mothers were absent due to work, illness, hospitalisation, or death may have differed systematically in nutritional status. Excluding these groups could have influenced prevalence estimates and the observed associations (52).

Despite these limitations, this study has important strengths. It uses nationally representative BDHS data across three survey rounds over an eight-year period. To our knowledge, this is the first study to estimate MDD-8 using the latest nationally representative data for Bangladesh and to examine temporal trends. These findings provide timely and policy-relevant evidence for monitoring progress in child feeding practices.

### 4.2 Future research

Future research should consider longitudinal designs to better understand causal pathways linking maternal, household, and service-related factors with dietary diversity. Qualitative research is also needed to explore caregivers’ perspectives on cultural beliefs, social norms, and practical barriers to providing diverse diets. Further studies could examine the effects of COVID-19, inflation, climate shocks, and conflict-related disruptions on child dietary diversity in Bangladesh. Additional work is also needed to explore the links between dietary diversity and child malnutrition outcomes, the reasons for higher dietary diversity in Chattogram and Sylhet, and the cost of achieving MDD-8 across wealth groups to inform subsidies, cash transfers, or food voucher programmes.

## 5. Conclusion

Dietary diversity among Bangladeshi children aged 6–23 months improved substantially between 2014 and 2017 but showed little progress between 2017 and 2022. Intake of several nutrient-dense food groups, including legumes and nuts, eggs, and fruits and vegetables, remained inadequate. Key determinants of MDD-8 included child age, birth order, maternal education, maternal employment, media exposure, ANC and PNC utilisation, household wealth, and region. Improving dietary diversity will require a multisectoral response that strengthens maternal education, women’s empowerment, access to maternal health services, affordability of diverse foods, targeted behaviour change communication, and support for disadvantaged areas. Addressing these gaps is critical for achieving SDG targets and improving child nutrition in Bangladesh.

## Data Availability

All data produced are available online at https://dhsprogram.com

https://dhsprogram.com

## Acknowledgements

We would like to express our gratitude to the Demographic Health Survey (DHS) Program for providing data access used in this research.

## Data Availability Statement

Publicly available data were used, which was accessible from the DHS website (https://dhsprogram.com) upon request.

## Authorship

IM, MAM, KTR, and TH conceived the study. IM conducted data curation and formal analysis. IM, MAM, KTR, and TH contributed to the methodology. IM wrote the original draft. IM worked on data visualisation. TH supervised the whole study. All authors contributed to the manuscript revision. All authors read and approved the final draft.

